# Inequalities in access to health care: an exploration of the cascade of care for in-transit migrants and asylum seekers, and the general population in Mexico

**DOI:** 10.1101/2025.01.09.25320284

**Authors:** Arjun Banerjee, César Rodríguez-Chavez, Jaime Sepúlveda, Steffanie Strathdee, Ietza Bojorquez

## Abstract

**BACKGROUND:** All over the world, migrants experience inequalities in access to health care. While this issue has been amply explored among immigrants and refugees in high-income countries, there is a dart of information on the situation of in-transit migrants and asylum seekers (IMAS) in low- or middle-income countries. Since these are the main recipient countries for this population, it is important to document inequalities in access to care in this context, especially for countries such as Mexico, in which IMAS are entitled to health care in par with the general population.

**OBJECTIVE:** to assess the inequalities in access to health care experienced by in-transit migrants in Mexico, as compared to the general population of Mexico, employing a cascade of care approach.

**METHODS:** We analyzed data from a cross-sectional survey of IMAS in shelters in Tijuana (November 2020-April 2021, n=481), and from ENSANUT-COVID 2020, a representative survey of the general population in Mexico conducted at about the same time (unweighted n=25,959). We compared the cascade of care – the self-reported presence of health needs, whether participants sought care given the need, and whether they received care – of the two samples.

**RESULTS:** A higher percentage of IMAS had experienced a health need in the past three months, as compared to the general population of Mexico (43.0% vs 21.9%). IMAS were less likely to seek care given the need (58.9% vs 81.8%), to receive care when they sought it (86.9% vs 97.7%), and to receive care in the public health system (19.7% vs 43.3%).

**CONCLUSIONS:** Inequalities in access to care were apparent, even in a health system that normatively should provide care regardless of migration status. Understanding why this occurs is important, and our results represent a baseline from which to evaluate future improvements in universal health coverage in Mexico.

## INTRODUCTION

Access to health care for migrants is an important component of universal health coverage, a global priority in WHO Sustainable Development Goals (Ghebreyesus, 2017; Onarheim et al., 2018). However, all over the world migrants face additional barriers for accessing health care, when compared to local populations (Abubakar et al., 2018; Bojorquez et al., 2021). On the other hand, while economic migrants may have better health outcomes than the local population (the healthy migrant effect) (Markides & Rote, 2019), this is not necessarily the case for in-transit migrants and asylum seekers (IMAS) who are forced out of their countries of origin by environmental, economic, political, or social crises (Pierola & Rodríguez Chatruc, 2020). These migrants can have poorer health outcomes compared to the general population with regards to communicable disease, noncommunicable disease, and mental health problems, due to various factors, such as the prevalence of communicable diseases in their countries of origin, poor living conditions while in transit, and the physical harms and psychological stresses associated with travel and migration (Leyva-Flores et al., 2019).

At the same time that the conditions of transit may increase health risks, they can further limit access to care, especially in low- and middle-income countries with poorly resourced health systems (Bojorquez et al., 2021). Among the issues that limit the access of IMAS to health care in this context, legal barriers, affordability issues, lack of information regarding access to services, language and cultural barriers, and discrimination have been documented (Ben Farhat et al., 2018; Bojorquez et al., 2021; Infante, Vieitez-Martinez, et al., 2022; Stevenson et al., 2023).

Mexico is a main transit country, with hundreds of thousands of IMAS travelling through it each year, most of them aiming to reach the United States (Rodriguez, 2016). Restrictive migration policies, both before and after the COVID-19 pandemic, including the 2018 “Remain in Mexico” order, the July 2019 “Third Country Asylum” rule, the use of Title 42 regulations during the pandemic, and the 2024 Executive Orders curtailing asylum claims at the Mexico-United States border, make thousands of IMAS wait at the Mexico-US border for extended periods, living in shelters or other informal accommodations, while waiting to continue their migration journey.

During this period of interrupted transit, health care for IMAS should be covered by the Mexican public health system, as the Mexican law grants access to universal health care, regardless of immigration status (Secretaría de Salud, 2019). This is in contrast with the situation in other regions, that exclude migrants from care or limit it to emergencies, as in the case of several European Union nations (Casquilho-Martins & Ferreira, 2022; Norredam et al., 2006), but is more typical of Latin America, where many countries have universal health systems that extend access to care to all types of migrants (Bojorquez, Cubillos-Novella, et al., 2024; Pierola & Rodríguez Chatruc, 2020). However, studies of IMAS in Mexico have identified that administrative barriers - such as requesting identity documents -, and the time and financial impact of transport to healthcare services can act as deterrents to accessing care (Infante, Vieitez-Martinez, et al., 2022; Leyva et al., 2016).

Despite the relevance of these issues, most studies of migrant’s access to health care are conducted in high-income countries, where immigrants or refugees and asylum seekers have already arrived at their intended destination. It is therefore necessary to increase the evidence base on access to health care of IMAS, with a focus on health inequalities between them and the general population of the countries they travel through. This is particularly important since low- and middle-income countries are the main recipients of IMAS (UNHCR, 2024). It is also important to assess in what stages of the process of access the inequalities are more marked, in order to better understand the mechanisms that limit equality.

Our aim in this study was to assess the inequalities in access to health care experienced by IMAS, as compared to the general population of Mexico, employing a cascade of care approach.

## METHODS

### Design and participants

We analyzed cross-sectional data from two surveys, one focused on IMAS, and the other a national health survey in Mexico, conducted at approximately the same time.

Data on IMAS were collected as part of a project that aimed to assess the impacts of the COVID-19 pandemic on migrants in Tijuana (n=481). The methods have been detailed previously (Bojorquez 2022). Briefly, a nonprobability sample of IMAS living in Tijuana, a city on the Mexico-United States border, was interviewed in November 2020-April 2021 (with an interruption in December 2020-January 2021 because of health regulations concerning COVID-19). Inclusion criteria were: (a) either having been born in a Latin American or Caribbean country other than Mexico, and having been in Tijuana for ⩽5 years, or having been born in a Mexican state other than Baja California, and having been in Tijuana for ⩽1 year due to deportation from the United States or internal displacement; (b) being ⩾18 years old; and (c) being able to respond the survey questionnaire in Spanish, English, or French. The sample size was calculated to estimate the prevalence of COVID-19 in this population. As reported previously, the statistical representativeness of the sample is unknown, but since data collection was conducted in the majority of the city’s migrant shelters, it is likely that the results accurately reflect IMAS in this context.

On the other hand, data on the general population was obtained from the open access datasets of ENSANUT, a probability household survey that is representative of the Mexican population. Specifically, we employed data from the ENSANUT-COVID 2020. The methods and other information of ENSANUT can be consulted at https://ensanut.insp.mx/index.php. We analyzed data from adults from 18 years old (unweighted n=25,959).

### Measures

All variables were measured through interviewer-administered questionnaires that included sociodemographic, migration, and health-related questions. The questionnaires were administered by previously trained interviewers on electronic devices. The research team that designed the IMAS survey took questions regarding health needs and access to care from ENSANUT questionnaire, so the responses are comparable between the two surveys.

The cascade of care approach, originally developed for analyses of the continuum of care in HIV, has subsequently been employed for other health issues (Colchero et al., 2023; Socias et al., 2016). It examines the stages in the process of access to care, measuring what proportion of those with a health need seek and obtain care. We employed the questions: “In the past 3 months, have you had any health needs?”, “did you seek care for that need?” and “did you receive care for that need?” (the latter two asked in regard to the most recent health need) to construct the cascade of care.

Furthermore, those who reported not seeking care for their most recent health need were asked the reason, with multiple response options. After reviewing frequencies of responses in both the IMAS survey and ENSANUT, we combined them into the following categories: thought the problem was unimportant, financial constraints, afraid of getting COVID-19 when going to a health care facility, and time constraints. Two additional options that were only asked in the IMAS survey: afraid of being identified as a migrant, and didn’t know where two seek care.

For descriptive purposes, we classified the type of need using a close-ended question into acute infectious (e.g. acute respiratory disease), chronic (e.g. diabetes), preventive (e.g. vaccination), acute noninfectious (e.g. accidents), mental health (e.g. depression), and other. The complete list of health needs in each category can be consulted in the ENSANUT documentation.

Finally, we classified the type of service were participants had received care as public (institutions that are part of social security systems, as well as care provided directly by the Ministry of Health), private, pharmacy-adjacent doctor’s office, Civil Society Organization (CSO)/charity services, and other.

### Analysis

Following a similar approach to Colchero et al. (2023), for the cascade of care we describe the percentages in each sample that: a) reported having a health need in the past 3 months; b) reported seeking care for their more recent health need; c) reported receiving care for their more recent health need; and d) reported receiving care for their more recent health need in the public system.

We considered the differences in these percentages (weighted estimators in the case of ENSANUT) between IMAS and the general population in Mexico as indicators of inequalities. In addition, we report confidence intervals for the ENSANUT estimators, allowing us to assess if the IMAS survey percentages fall within the intervals’ limits.

Since the main sociodemographic characteristics of the two samples differ, we accounted for those differences by sensitivity analyses restricted by gender (female), age group (40+ years), and education level (<=9 years of education), the later as a proxy for socioeconomic level.

Statistical analysis was conducted with Stata 17, using the svy module when appropriate (StataCorp 2024).

### Ethical Considerations

The IMAS survey was approved by the ethical review committees of El Colegio de la Frontera Norte (no. 066-170720) and the University of California, San Diego (no. 201153). Participants provided written informed consent in Spanish, French or English. The ENSANUT was approved by the Ethics Committee of the National Institute of Public Health, Mexico, and all participants provided informed consent.

## RESULTS

The IMAS study sample included 481 IMAS living in shelters. Slightly over half (55.3%) were women, most were 18-39 years of age (77.6%), and the majority had an educational level of nine years or less (62.5%). In comparison, the percentage of women in the adult population in Mexico (unweighted n=25,959) was lower (51.9%), the population was older (50.4% were 40 years old or over), and the educational level was higher (52.8% with nine years or less of education) (Table 1).

**Table 1.**
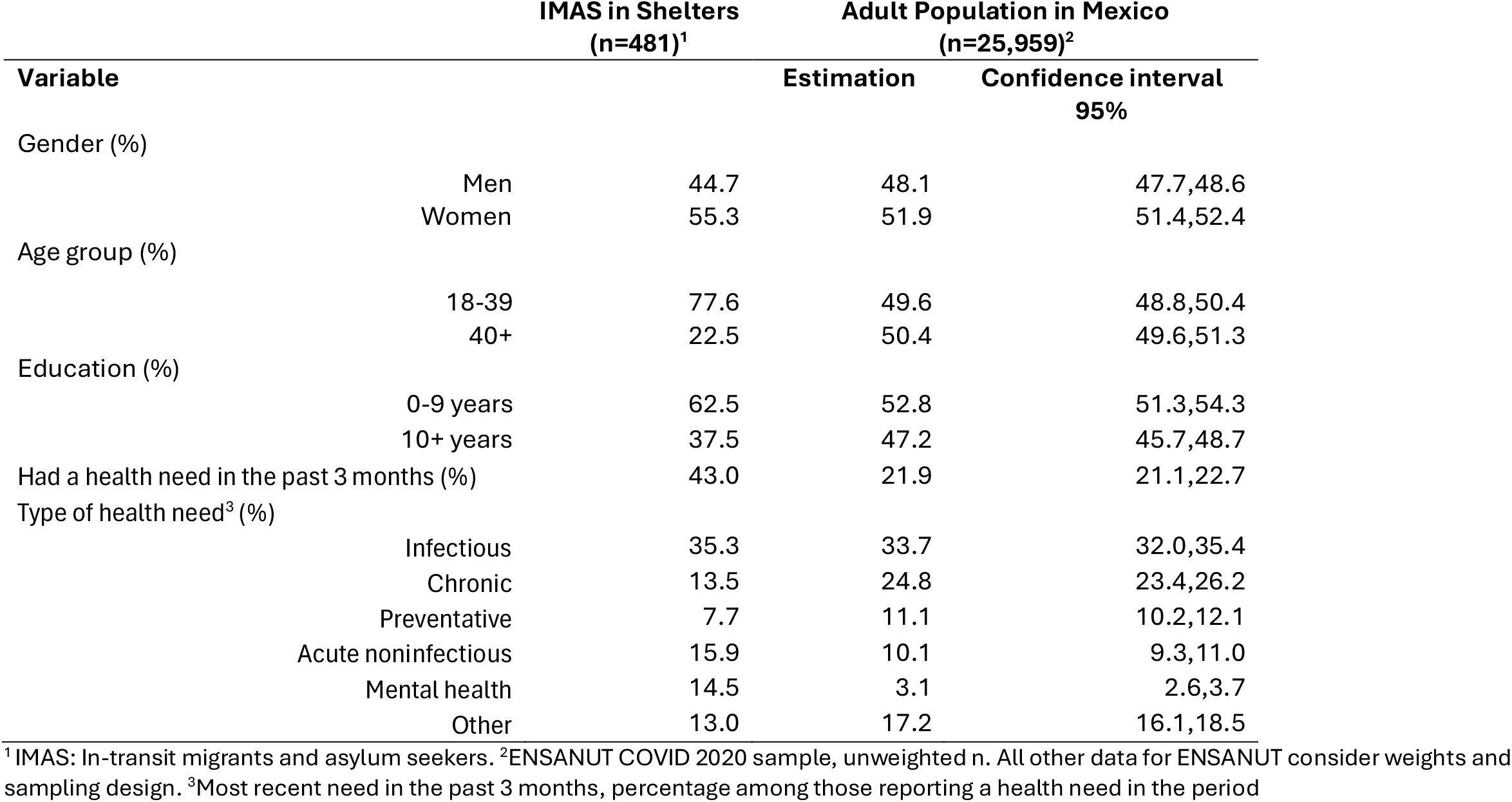
Characteristics of participants.

IMAS were more likely to report a health need in the past three months than the adult population in Mexico (43.0% vs 21.9%) (Table 1 & Figure 1). The most frequent type of health need in both groups were infectious diseases (35.3% and 33.7%, respectively). The second most frequent type of need among IMAS, more frequent in this group than in the adult population in Mexico, were acute noninfectious needs (15.9% vs 10.1% in ENSANUT), mainly due to reports of headache of unspecific cause or accidents (not shown in Table). The third more frequent health needs for IMAS were mental health-related (14.5%), which were unfrequently mentioned by ENSANUT participants (3.1%).

**Figure 1.**
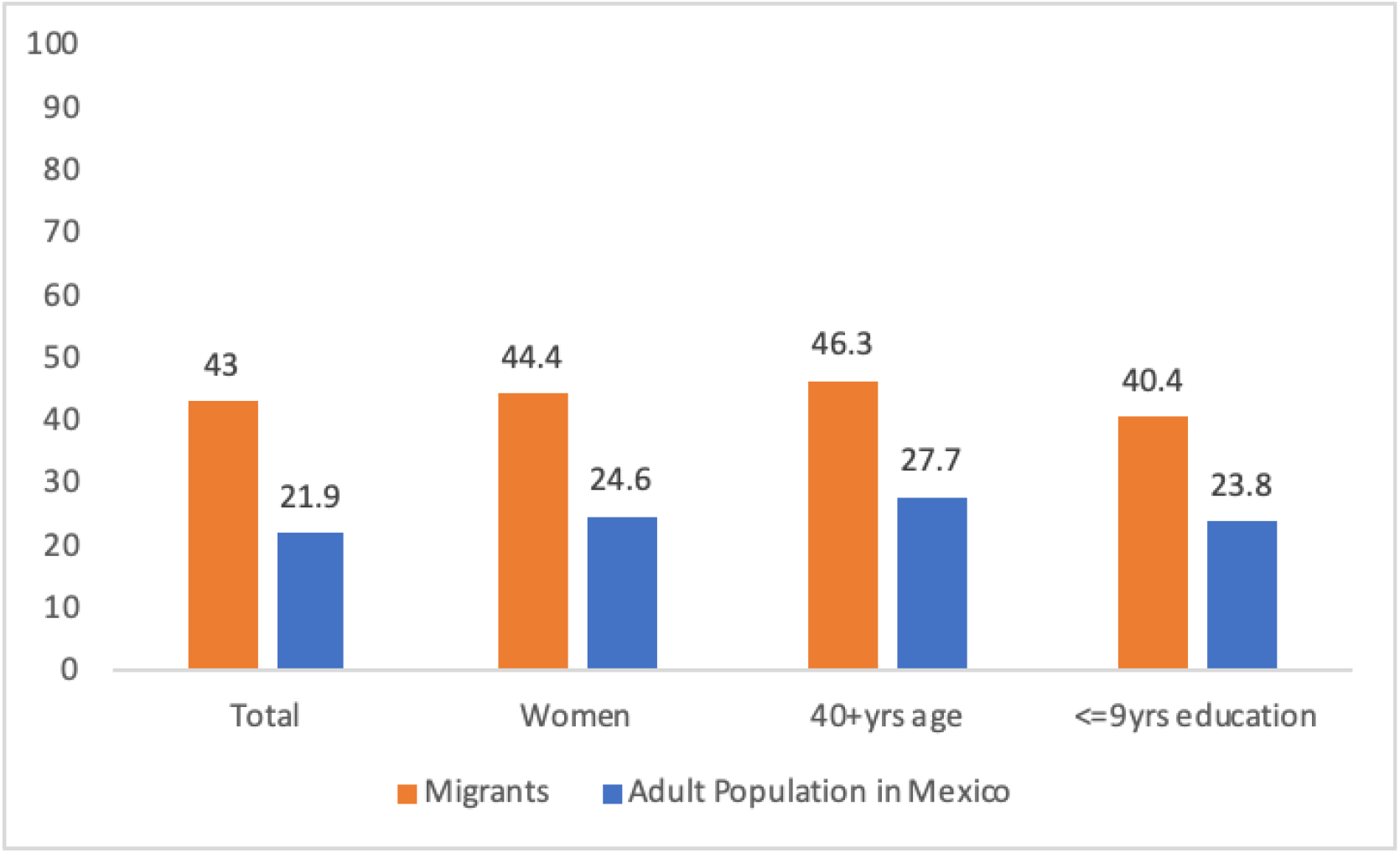
Percentage with a health need in the past three months, IMAS and adult population in Mexico, by sociodemographic characteristics^1^. ^1^ IMAS: In-transit migrants and asylum seekers. Adult population in Mexico from the ENSANUT COVID 2020 sample

Of those reporting a health need in the past three months, 58.9% of IMAS and 81.8% of the adult population in Mexico sought care for that need (Table 2). Of those who sought care, 86.9% of IMAS and 97.7% of the ENSANUT sample received it, and of those who received care, 19.7% of IMAS and 43.3% of the ENSANUT sample received it in a public facility. From these data, the relative inequalities between IMAS and the adult population in Mexico were 0.72 for care seeking given the need (IMAS were 72% as likely to seek care as the ENSANUT sample), 0.89 for receiving care given it was sought, and 0.45 for receiving care in a public facility given it was received.

**Table 2.** Access to health care.

|  | IMAS in Shelters<br>with a health need<br>in the past 3<br>months<br>(n=207)<sup>1</sup> | Adult Population in Mexico with a<br>health need in the past 3 months<br>(n=5,954)<sup>2</sup> |  |
| --- | --- | --- | --- |
| Variable |  | Estimation | Confidence<br>interval 95% |
| **Cascade of care** |  |  |  |
| % sought care (out of those with a health need) | 58.9 | 81.8 | 80.4,83.1 |
| % received care (out of those who sought care) | 86.9 | 97.7 | 97.2,98.1 |
| **Reasons care was not sought<sup>3</sup>** |  |  |  |
| thought the problem wasn't important | 20.0 | 51.6 | 47.8,55.4 |
| financial constraints | 55.3 | 15.0 | 12.3,18.2 |
| afraid of getting COVID-19 | 5.9 | 13.3 | 11.0,15.9 |
| time constraints | 3.5 | 8.6 | 6.8,10.7 |
| afraid of being identified as a migrant | 1.2 | n.a.<sup>4</sup> |  |
| didn't know where to seek care | 29.4 | n.a.<sup>4</sup> |  |
| **Where care was sought<sup>5</sup>** |  |  |  |
| public | 19.7 | 43.3 | 41.5,45.2 |
| private | 9.0 | 37.8 | 35.9,39.8 |
| pharmacy-adjacent | 19.7 | 16.5 | 15.1,18.0 |
| CSO, charity | 31.2 | 0.3 | 0.2,0.6 |
| other | 20.5 | 2.0 | 1.6,2.6 |

<sup>1</sup> IMAS: In-transit migrants and asylum seekers. <sup>2</sup> ENSANUT COVID 2020 sample, unweighted n. All other data for ENSANUT consider weight and sampling design. <sup>3</sup> Multiple responses allowed. <sup>4</sup> Not applicable (option not available in the ENSANUT questionnaire). <sup>5</sup> Percentage out of those who received care

These differences are shown in a slightly different way in Figure 2, where the percentages of those who received care, and of those who received care in a public facility, are calculated over the total reporting a health need in the past three months. In contrast with the data in Table 2, Figure 2 thus presents a comparison in the endpoint of access for this analysis: receiving care in a public facility, in a country with a universal health care system that is responsible for the care of all persons. For this endpoint, IMAS were only 34% as likely as the ENSANUT sample to receive care in a public facility, given a health need (11.6% vs 34.6%). For migrants with nine years or less of education, the difference was even more pronounced: IMAS were only 28% as likely as the equivalent ENSANUT sample to receive care in a public facility given a health need (10.4% vs 37.2%).

**Figure 2.**
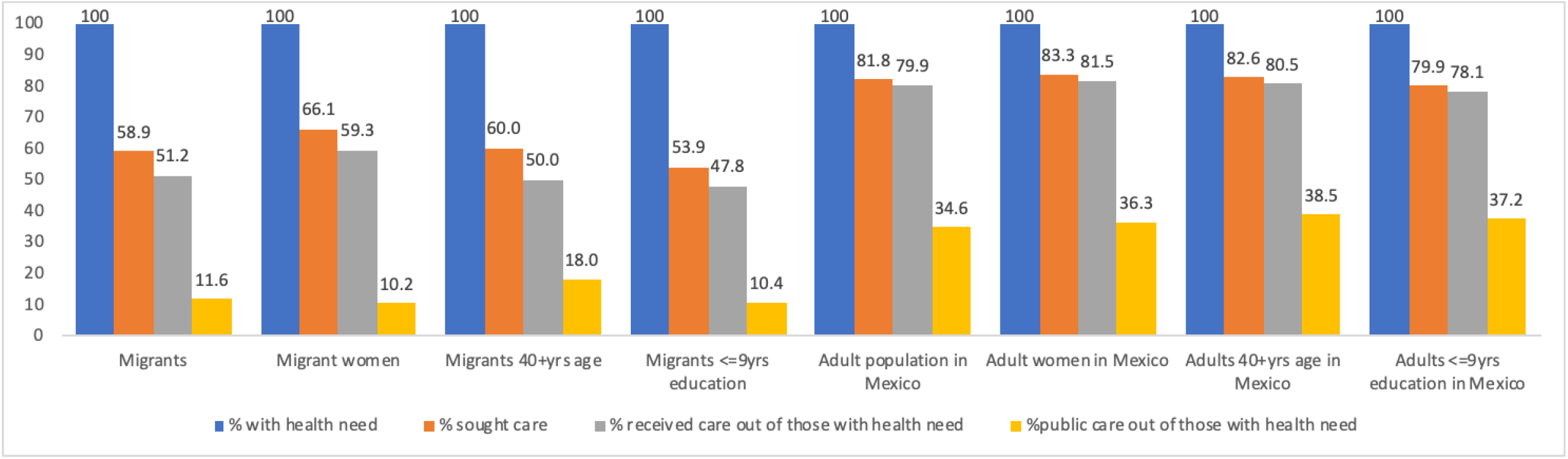
Cascade of care, IMAS and adult population in Mexico, by sociodemographic characteristics ^1^. ^1^ IMAS: In-transit migrants and asylum seekers. Adult population in Mexico from the ENSANUT COVID 2020 sample

On the other hand, figures 1 & 2 (and the Supplementary Tables) show that the differences observed persisted after restricting the analysis by gender, age group, and education level. These variables are highly collinear, and the small sample size for some categories of IMAS limits the interpretation of the results. However, the patterns are similar to those in the whole dataset, with inequalities affecting IMAS in terms of both health needs and access to care.

Finally, IMAS were more likely to report forgoing seeking treatment due to lack of money than the adult Mexican population (55.3% vs 15.0%). The second most common reason for not seeking care reported by IMAS was not knowing where to go for it (29.4%), and the third one thinking the health need was not important (20.0%). In contrast, the three main reasons for not seeking care among the adult Mexican population, in order of importance, were thinking the health need was not important (51.6%), financial constraints (15.0%), and fear of getting COVID-19 (13.3%) (Table 1).

## DISCUSSION

The aim of our study was to compare the cascade of care of IMAS and the adult population in Mexico, in order to assess inequalities in access to care between the two groups, and the stages in the process of access where these inequalities occur. Our results show that, even while a higher proportion of IMAS reported having had a health need recently, they were less likely to seek and access care, and to seek care in the public system. These results are concerning, considering that Mexico is a country where all persons, regardless of their migration status, are entitled to health care (Secretaría de Salud, 2019).

Inequalities were evident since the first step of care seeking, as only 58.9% of IMAS sought care when faced with a health need, as compared to 81.8% of the general population. The reasons they gave for not seeking care (financial constraints and lack of information) should be relatively easy to address. As for the former, providing care for the main health needs of IMAS in transit through Mexico would require only a small percentage of the country’s health budget (Bojorquez, Cerecero, et al., 2024). As for the latter, and organizing campaigns to inform IMAS of their right to health care and how to access the public system should be straightforward.

On the other hand, the main relative inequality between IMAS and the adult population in Mexico occurred in the utilization of public health facilities. This is extremely important, since the expectation in this system is that care is provided without the need for out of pocket expenditures. As IMAS mentioned financial concerns as their number one reason for not seeking care, it is important to further investigate the motives why they did not resort to this, supposedly free system. One likely reason is that care in the public system in Mexico is not completely free, as patients frequently have to purchase medicines or pay for lab tests that are unavailable in the public facilities (Gonzalez Block et al., 2020). A second, related one, is that the health needs of IMAS are cared for by CSOs, charities and volunteers that provide services (including medicines) free of charge. As reported previously (Infante, Bojorquez, et al., 2022; Leyva et al., 2016) these organizations were the main source of care for IMAS in our study.

Finally, while this article is not focused on the prevalence of specific health care needs, it is noteworthy that IMAS were significantly more likely to report mental health issues than the general Mexican population. The high prevalence of mental health problems including depression and anxiety among IMAS in Mexico has been previously reported (Bojorquez et al., 2022; Bojorquez-Chapela et al., 2023), and our work further shows the need to direct resources to mental health programs and interventions

A limitation of our study is the relatively small, non-probability sample of IMAS. While we believe it is necessary to improve the quality and representativeness of studies of IMAS, data on this population is difficult to collect, and our study adds to the documentation of the health-related challenges faced by this population. On the other hand, in order to assess the disparities that affect IMAS, it is necessary to make comparisons between them and non-migrant populations. This type of research is incipient in Latin America, and almost non-existent in regard to IMAS, so we believe our work provides a meaningful starting point for future studies.

Another important caveat when interpreting our study is that the data were collected at the peak of the COVID-19 pandemic. During that time, migrants bore the brunt of the public health emergencies compounded by their already vulnerable situation (Cubillos Novella et al., 2023; Stevenson et al., 2023), and similar studies with data from a different time may render different results. However, the inequities in healthcare access among IMAS in this study reflect those that migrants face worldwide (Abubakar et al., 2018; Busetta et al., 2018).

In conclusion, our work identifies significant inequalities throughout the process of access to health care between IMAS and the general adult population in Mexico. Our study identified affordability of care and lack of information as two main reasons for not seeking care, pointing to aspects that could be addressed by public policy. It is important to continue exploring the process of access to health care for IMAS, in order to document the current situation, and to monitor changes in the road to universal health care.

## Funding

This work was supported by the American Academy of Arts and Sciences’ Rethinking the Humanitarian Health Response to Violent Conflict Project [no grant number]; the University of California, San Francisco [no grant number]; and the San Diego Center For AIDS Research (CFAR), which is funded by the National Institutes of Health [grant number P30AI036214].

## Conflict of interest

The authors declare that they had no conflict of interest

## Data availability

Data from the IMAS study are available upon reasonable request from the corresponding author. ENSANUT is publicly available at https://ensanut.insp.mx/

## CRediT author statement

AB: Analysis; writing-original draft; writing-review & editing. CRC: Writing-original draft; writing-review & editing. JS: Conceptualization and conduction of the IMAS survey; writing-review & editing; supervision; funding acquisition. SS: Conceptualization and conduction of the IMAS survey; writing-review & editing; supervision; funding acquisition. IB: Conceptualization and conduction of the IMAS survey; conceptualization of the analysis for this article; methodology; analysis; writing-original draft; writing-review & editing; visualization; supervision

## Notes

### Competing Interest Statement

The authors have declared no competing interest.

### Funding Statement

The IMAS study was funded by the American Academy of Arts and Sciences' Rethinking the Humanitarian Health Response to Violent Conflict Project [no grant number]; the University of California, San Francisco [no grant number]; and the San Diego Center For AIDS Research (CFAR), which is funded by the National Institutes of Health [grant number P30AI036214].

### Author Declarations

The Ethics Committees of El Colegio de la Frontera Norte (no. 066-170720) and the University of California, San Diego (no. 201153) gave ethical approval for this work

## References

Abubakar, I., Aldridge, R. W., Devakumar, D., Orcutt, M., Burns, R., Barreto, M. L., Dhavan, P., Fouad, F. M., Groce, N., Guo, Y., Hargreaves, S., Knipper, M., Miranda, J. J., Madise, N., Kumar, B., Mosca, D., McGovern, T., Rubenstein, L., Sammonds, P., Sawyer, S. M., Sheikh, K., Tollman, S., Spiegel, P., Zimmerman, C., & U. CL-Lancet Commission on Migration and Health. (2018, Dec 15). The UCL-Lancet Commission on Migration and Health: the health of a world on the move. Lancet, 392(10164), 2606–2654. 10.1016/S0140-6736(18)32114-7

Ben Farhat, J., Blanchet, K., Juul Bjertrup, P., Veizis, A., Perrin, C., Coulborn, R. M., Mayaud, P., & Cohuet, S. (2018, Mar 13). Syrian refugees in Greece: experience with violence, mental health status, and access to information during the journey and while in Greece. BMC Med, 16(1), 40. 10.1186/s12916-018-1028-4

Bojorquez, I., Cabieses, B., Arosquipa, C., Arroyo, J., Novella, A. C., Knipper, M., Orcutt, M., Sedas, A. C., & Rojas, K. (2021, Apr 3). Migration and health in Latin America during the COVID-19 pandemic and beyond. Lancet, 397(10281), 1243–1245. 10.1016/S0140-6736(21)00629-2

Bojorquez, I., Cerecero, D., Orraca-Romero, P., Fernandez-Niño, J. A., Rojas-Botero, M. L., & Infante, C. (2024). Atención en salud a migrantes en tránsito: estimación del costo para el sistema de salud en México. Salud Publica de Mexico, 66, 816–823. 10.21149/15736

Bojorquez, I., Cubillos-Novella, A., Arroyo-Laguna, J., Martinez-Juarez, L. A., Sedas, A. C., Franco-Suarez, O., Suárez-Morales, Z., Adame-Avilés, E., Barragán-León, M., Suarez, A., Orcutt, M., & Spiegel, P. (2024). The response of health systems to the needs of migrants and refugees in the COVID-19 pandemic: a comparative case study between Mexico, Colombia and Peru. The Lancet Regional Health – Americas, 40, 100763. 10.1016/j.lana.2024.100763 ISSN 2667-193X

Bojorquez, I., Sepulveda, J., Lee, D., & Strathdee, S. (2022, Jun 2). Interrupted transit and common mental disorders among migrants in Tijuana, Mexico. International Journal of Social Psychiatry, 207640221099419. 10.1177/00207640221099419

Bojorquez-Chapela, I., Leyva-Flores, R., Gómez-López, D., Vazquez, E. K., & Cortes-Alcala, R. (2023). Forced migration and psychological distress among migrants in transit through Mexico. Salud Publica de Mexico, 66(2), 173–181. 10.21149/14829

Busetta, A., Cetorelli, V., & Wilson, B. (2018, Apr). A Universal Health Care System? Unmet Need for Medical Care Among Regular and Irregular Immigrants in Italy. Journal of Immigrant and Minority Health, 20(2), 416–421. 10.1007/s10903-017-0566-8

Casquilho-Martins, I., & Ferreira, S. (2022). Migrants’ Health Policies and Access to Health Care in Portugal within the European Framework. Societies, 12(2), 55. https://www.mdpi.com/2075-4698/12/2/55

Colchero, M. A., Gomez, R., & Bautista-Arredondo, S. (2023, Jan 2). Elección de proveedores de servicios de atención y necesidades de salud de la población mexicana, 2021. Salud Publica de Mexico, 65(1), 28–35. 10.21149/14107

Cubillos Novella, A., Eguren, J., & Ochoa Marín, C. (Eds.). (2023). Los desafíos de la migración a la salud pública en Iberoamérica en tiempos de la COVID-19. Pontificia Universidad Javeriana.

Ghebreyesus, T. A. (2017, Sep). All roads lead to universal health coverage. Lancet Glob Health, 5(9), e839–e840. 10.1016/S2214-109X(17)30295-4

Gonzalez Block, M. A., Reyes Morales, H., Hurtado, L. C., Balandran, A., & Mendez, E. (2020, Apr). Mexico: Health System Review. Health Syst Transit, 22(2), 1–222. https://www.ncbi.nlm.nih.gov/pubmed/33527902

Infante, C., Bojorquez, I., Vieitez-Martinez, I., Larrea-Schiavon, S., Napoles-Mendez, G., & Rodriguez-Chavez, C. (2022). Migrant shelters’ response to COVID-19: Comparative case study in four cities close to the Mexico-United States border. J Migr Health, 6, 100110. 10.1016/j.jmh.2022.100110

Infante, C., Vieitez-Martinez, I., Rodriguez-Chavez, C., Napoles, G., Larrea-Schiavon, S., & Bojorquez, I. (2022). Access to Health Care for Migrants Along the Mexico-United States Border: Applying a Framework to Assess Barriers to Care in Mexico. Front Public Health, 10, 921417. 10.3389/fpubh.2022.921417

Leyva, R., Infante, C., & Quintino, F. (2016). Migrantes en tránsito por México: Situación de salud, riesgos y acceso a servicios de salud. INSP.

Leyva-Flores, R., Infante, C., Gutierrez, J. P., Quintino-Perez, F., Gomez-Saldivar, M., & Torres-Robles, C. (2019). Migrants in transit through Mexico to the US: Experiences with violence and related factors, 2009-2015. PLoS One, 14(8), e0220775. 10.1371/journal.pone.0220775

Markides, K. S., & Rote, S. (2019, Mar 14). The Healthy Immigrant Effect and Aging in the United States and Other Western Countries. Gerontologist, 59(2), 205–214. 10.1093/geront/gny136

Norredam, M., Mygind, A., & Krasnik, A. (2006, Jun). Access to health care for asylum seekers in the European Union--a comparative study of country policies. Eur J Public Health, 16(3), 286–290. 10.1093/eurpub/cki191

Onarheim, K. H., Melberg, A., Meier, B. M., & Miljeteig, I. (2018). Towards universal health coverage: including undocumented migrants. BMJ Global Health, 3(5), e001031. 10.1136/bmjgh-2018-001031

Pierola, M. D., & Rodríguez Chatruc, M. (2020). Migrants in Latin America: Disparities in Health Status and in Access to Healthcare. I.D. Bank. 10.18235/0002432

Rodriguez, E. (2016). Migración centroamericana en tránsito irregular por México: nuevas cifras y tendencias. CIESAS.

Secretaría de Salud. (2019, 29 de Noviembre de 2019). Decreto por el que se reforman, adicionan y derogan diversas disposiciones de la Ley General de Salud y de la Ley de los Institutos Nacionales de Salud. Diario Oficial de la Federación, 101–117.

Socias, M. E., Volkow, N., & Wood, E. (2016, Dec). Adopting the ‘cascade of care’ framework: an opportunity to close the implementation gap in addiction care? Addiction, 111(12), 2079–2081. 10.1111/add.13479

Stevenson, M., Guillén, J. R., Bevilacqua, K. G., Arciniegas, S., Ortíz, J., López, J. J., Ramírez, J. F., Talero, M. B., Quijano, C., Vela, A., Moreno, Y., Rigual, F., Page, K. R., Spiegel, P. B., Núñez, R. L., Fernández-Niño, J. A., & Wirtz, A. L. (2023, 2023/01/01/). Qualitative assessment of the impacts of the COVID-19 pandemic on migration, access to healthcare, and social wellbeing among Venezuelan migrants and refugees in Colombia. Journal of Migration and Health, 7, 100187. 10.1016/j.jmh.2023.100187

UNHCR. (2024). Global Trends: Forced Displacement in 2023. https://www.unhcr.org/refugee-statistics

