## Supplementary Tables for "Inequalities in access to health care: an exploration of the cascade of care for in-transit migrants and asylum seekers, and the general population in Mexico"

**Table 1a. Characteristics of participants (women)**

| | IMAS in Shelters (n=266)[1] | Adult Population in Mexico (n=13,812)[2] | |
|---|---|---|---|
| **Variable** | | **Estimation** | **Confidence interval 95%** |
| Age group | | | |
| 18-39 | 86.1 | 48.3 | 47.4,49.3 |
| 40+ | 13.9 | 51.7 | 50.7,52.7 |
| Education | | | |
| 0-9 years | 62.9 | 53.6 | 52.0,55.2 |
| 10+ years | 37.2 | 46.4 | 44.8,48.0 |
| Had a health need in the past 3 months (%) | 44.4 | 24.6 | 23.7,25.6 |
| Type of health need[3] (%) | | | |
| Infectious | 34.8 | 31.8 | 29.8,33.8 |
| Chronic | 11.9 | 25.7 | 24.0,27.5 |
| Preventative | 11.9 | 12.6 | 11.3,13.9 |
| Acute noninfectious | 14.4 | 9.0 | 8.0,10.1 |
| Mental health | 15.3 | 3.5 | 2.8,4.3 |
| Other | 11.9 | 17.5 | 16.0,19.2 |

[1] IMAS: In-transit migrants and asylum seekers. [2]ENSANUT COVID 2020 sample, unweighted n. All other data for ENSANUT consider weights and sampling design. [3]Most recent need in the past 3 months, percentage among those reporting a health need in the period

**Table 1b. Characteristics of participants (40 years of age or older)**

| | IMAS in Shelters (n=108)[1] | Adult Population in Mexico (n=14,557)[2] | |
|---|---|---|---|
| **Variable** | | **Estimation** | **Confidence interval 95%** |
| Gender (%) | | | |
| Male | 65.7 | 46.8 | 46.2,47.4 |
| Female | 34.3 | 53.2 | 52.6,53.8 |
| Education | | | |
| 0-9 years | 65.7 | 67.3 | 65.4,69.2 |
| 10+ years | 34.3 | 32.7 | 30.9,34.6 |
| Had a health need in the past 3 months (%) | 46.3 | 27.7 | 26.6,28.8 |
| Type of health need[3] (%) | | | |
| Infectious | 28.0 | 26.2 | 24.4,28.0 |
| Chronic | 24.0 | 32.5 | 30.7,34.3 |
| Preventative | 8.0 | 11.9 | 10.7,13.1 |
| Acute noninfectious | 14.0 | 8.3 | 7.4,9.2 |
| Mental health | 14.0 | 3.0 | 2.5,3.6 |
| Other | 12.0 | 18.2 | 16.9,19.5 |

[1] IMAS: In-transit migrants and asylum seekers. [2]ENSANUT COVID 2020 sample, unweighted n. All other data for ENSANUT consider weights and sampling design. [2]Most recent need in the past 3 months, percentage among those reporting a health need in the period

**Table 1c. Characteristics of participants (nine years of education or less)**

| | IMAS in Shelters (n=285)[1] | Adult Population in Mexico (n=14,482)[2] | |
|---|---|---|---|
| Variable | | Estimation | Confidence interval 95% |
| Gender (%) | | | |
| Male | 44.2 | 47.3 | 46.6,48.0 |
| Female | 55.8 | 52.7 | 52.0,53.4 |
| Age group | | | |
| 18-39 | 77.2 | 35.9 | 34.6,37.2 |
| 40+ | 22.8 | 64.1 | 62.8,65.4 |
| Had a health need in the past 3 months (%) | 40.4 | 23.8 | 22.8,24.8 |
| Type of health need[3] (%) | | | |
| Infectious | 37.4 | 27.7 | 25.7,29.8 |
| Chronic | 13.9 | 29.9 | 28.0,31.8 |
| Preventative | 7.0 | 10.7 | 9.6,12.0 |
| Acute noninfectious | 11.3 | 10.8 | 9.7,11.9 |
| Mental health | 14.8 | 2.4 | 1.9,3.0 |
| Other | 15.7 | 18.5 | 17.0,20.1 |

[1] IMAS: In-transit migrants and asylum seekers. [2]ENSANUT COVID 2020 sample, unweighted n. All other data for ENSANUT consider weights and sampling design. [3]Most recent need in the past 3 months, percentage among those reporting a health need in the period

**Table 2a. Access to health care (women)**

| | IMAS in Shelters with a health need in the past 3 months (n=118)[1] | Adults in Mexico with a health need in the past 3 months (n=3,541)[2] | |
|---|---|---|---|
| Variable | | Estimation | Confidence interval 95% |
| **Cascade of care** | | | |
| % sought care (out of those with a health need) | 66.1 | 83.3 | 81.4,84.9 |
| % received care (out of those who sought care) | 89.7 | 97.8 | 97.2,98.3 |
| **Reasons care was not sought[3]** | | | |
| thought the problem wasn't important | 15.0 | 46.2 | 41.6,50.9 |
| financial constraints | 55.0 | 14.8 | 11.7,18.5 |
| I was afraid of getting COVID-19 | 5.0 | 16.9 | 13.6,20.9 |
| time constraints | 2.5 | 8.4 | 6.5,10.8 |
| afraid of being identified as a migrant | 0 | n.a.[4] | |
| didn't know where to seek care | 40.0 | n.a.[4] | |
| **Where care was sought[5]** | | | |
| public | 15.4 | 44.6 | 42.4,46.8 |
| private | 7.7 | 36.2 | 33.9,38.5 |
| pharmacy-adjacent | 16.7 | 16.7 | 15.0,18.6 |
| SCO, charity | 39.7 | 0.4 | 0.2,0.7 |
| other | 20.5 | 2.2 | 1.6,2.9 |

[1] IMAS: In-transit migrants and asylum seekers. [2]ENSANUT COVID 2020 sample, unweighted n. All other data for ENSANUT consider weight and sampling design.
[3] Multiple responses allowed. [4]Not applicable (option not available in the ENSANUT questionnaire). [5]Percentage out of those who received care

**Table 2b. Access to health care (40 years of age or older)**

| | IMAS in Shelters with a health need in the past 3 months (n= 50)[1] | Adult population Mexico with a health need in the past 3 months (n=4,137)[2] | |
|---|---|---|---|
| Variable | | Estimation | Confidence interval 95% |
| **Cascade of care** | | | |
| % sought care (out of those with a health need) | 60.0 | 82.6 | 81.1,84.0 |
| % received care (out of those who sought care) | 83.3 | 97.5 | 96.8,98.0 |
| **Reasons care was not sought[3]** | | | |
| thought the problem wasn't important | 5.0 | 44.2 | 39.9,48.5 |
| financial constraints | 65.0 | 14.8 | 11.7,18.5 |
| I was afraid of getting COVID-19 | 10.0 | 16.5 | 13.5,19.9 |
| time constraints | 5.0 | 9.4 | 7.5,11.8 |
| afraid of being identified as a migrant | 0 | n.a.[4] | |
| didn't know where to seek care | 10.0 | n.a.[4] | |
| **Where care was sought[5]** | | | |
| public | 30.0 | 47.9 | 45.6,50.2 |
| private | 0 | 36.4 | 34.3,38,6 |
| pharmacy-adjacent | 23.3 | 13.9 | 12.4,15.6 |
| SCO, charity | 33.3 | 0.3 | 0.1,0.5 |
| other | 13.3 | 1.6 | 1.2,2.1 |

[1] IMAS: In-transit migrants and asylum seekers. [2]ENSANUT COVID 2020 sample, unweighted n. All other data for ENSANUT consider weight and sampling design.
[3] Multiple responses allowed. [4]Not applicable (option not available in the ENSANUT questionnaire). [5]Percentage out of those who received care

**Table 2c. Access to health care (participants with nine years of education or less)**

| | IMAS in Shelters with a health need in the past 3 months (n=115)[1] | Adult Population in Mexico with a health need in the past 3 months (n=3,599)[2] | |
|---|---|---|---|
| Variable | | Estimation | Confidence interval 95% |
| **Cascade of care** | | | |
| % sought care (out of those with a health need) | 53.9 | 79.9 | 78.1,81.7 |
| % received care (out of those who sought care) | 88.7 | 97.7 | 96.9,98.2 |
| **Reasons care was not sought[3]** | | | |
| thought the problem wasn't important | 20.8 | 48.9 | 44.3,53.4 |
| financial constraints | 54.7 | 16.6 | 13.3,20.6 |
| I was afraid of getting COVID-19 | 7.6 | 14.2 | 11.6,17.3 |
| time constraints | 3.8 | 9.5 | 7.4,12.0 |
| afraid of being identified as a migrant | 0 | n.a.[4] | |
| didn't know where to seek care | 35.9 | n.a.[4] | |
| **Where care was sought[5]** | | | |
| public | 19.4 | 47.7 | 45.4,50.0 |
| private | 8.1 | 33.4 | 31.3,35.6 |
| pharmacy-adjacent | 19.4 | 16.8 | 15.0,18.8 |
| SCO, charity | 30.7 | 0.4 | 0.2,0.8 |
| other | 22.6 | 1.7 | 1.2,2.3 |

[1] IMAS: In-transit migrants and asylum seekers. [2]ENSANUT COVID 2020 sample, unweighted n. All other data for ENSANUT consider weight and sampling design.
[3] Multiple responses allowed. [4]Not applicable (option not available in the ENSANUT questionnaire). [5]Percentage out of those who received care
